# Development of criteria for cognitive dysfunction in post-COVID syndrome: the IC-CoDi-COVID approach

**DOI:** 10.1101/2022.10.21.22281239

**Authors:** Jordi A Matias-Guiu, Elena Herrera, María González-Nosti, Kamini Krishnan, Cristina Delgado-Álvarez, María Díez-Cirarda, Miguel Yus, Álvaro Martínez-Petit, Josué Pagán, Jorge Matías-Guiu, José Luis Ayala, Robyn Busch, Bruce P Hermann

**Author notes:** **Correspondence to:** Dr Jordi A Matias-Guiu. Department of Neurology, Hospital Clinico San Carlos. Prof. Martin Lagos St, 28040, Madrid (Spain).

## Abstract

**Background:** We aimed to develop objective criteria for cognitive dysfunction associated with the post-COVID syndrome.

**Methods:** Four hundred and four patients with post-COVID syndrome from two centers were evaluated with comprehensive neuropsychological batteries. The International Classification for Cognitive Disorders in Epilepsy (IC-CoDE) framework was adapted and implemented. A complementary data-driven approach based on unsupervised machine-learning clustering algorithms was also used to evaluate the optimal classification and cutoff points.

**Results:** According to the developed criteria, 41.2% and 17.3% of the sample were classified as having at least one cognitive domain impaired using -1 and -1.5 standard deviations as cutoff points. Attention/processing speed was the most frequently impaired domain. There were no differences in base rates of cognitive impairment between the two centers. Clustering analysis revealed two clusters according to the severity of cognitive impairment, but there was no difference in cognitive profiles. Cognitive impairment was associated with younger age and lower education levels, but not hospitalization.

**Conclusions:** We propose a harmonization of the criteria to define and classify cognitive impairment in the post-COVID syndrome. These criteria may be extrapolated to other neuropsychological batteries and settings, contributing to the diagnosis of cognitive deficits after COVID-19 and facilitating multicenter studies to guide biomarker investigation and therapies.

## 1. INTRODUCTION

SARS-CoV-2 infection has been associated with brain damage that can occur via a number of different mechanisms[1]. Importantly, these patients may report cognitive symptoms several weeks or months after the onset of COVID-19 [2-4]. Indeed, according to the World Health Organization (WHO) criteria for post-COVID syndrome, cognitive symptoms are among the most frequent symptoms of this new disorder[5]. Recent investigations have reported the broad characteristics of cognitive dysfunction associated with COVID-19, including impairments in attention, executive function, memory, visuospatial function, and language. The magnitude of cognitive deficits is generally small to moderate, and patients are often of working age[6-8]. Brief cognitive tests or screeners, such as those used in the dementia field, do not have adequate sensitivity to detect these deficits; therefore, comprehensive neuropsychological assessment is required.

Given the high frequency of COVID-19 and post-COVID syndrome, cognitive issues after COVID-19 have resulted in a marked increase in referrals to neurology and memory clinics[9]. Identification of cognitive dysfunction is important in clinical care for several reasons, including early treatment/enrollment in cognitive training/rehabilitation programs, participation in clinical trials, and job adaptation or disability benefits.

Different research groups investigating post-COVID syndrome or other neurological disorders potentially associated with cognitive impairment (e.g., epilepsy, multiple sclerosis) have characterized cognitive impairment and profiles using different criteria. For instance, individual tests may be classified as impaired or intact using a set cut-off (e.g., 1, 1.5, or 2 standard deviations below the normative mean) and specific cognitive domains may be classified based on impaired performance on one or two tests within a domain or when a specified percentage of the tests administered are below a given cutoff[7,10]. This variability makes it difficult to compare results across studies, and the use of heterogeneous assessment approaches, in which some tests or domains may be more represented than others, may bias the findings as well. In addition, the use of more conservative or liberal cutoffs may also produce false negatives or positives, respectively, in the diagnosis of cognitive dysfunction. Overall, these issues challenge the generalizability of the results across centers and countries and a fundamental understanding of the cognitive consequences of COVID-19.

To address these issues, it is important to develop objective neuropsychological criteria to define cognitive dysfunction in the context of COVID-19 as well as other conditions that affect cognition. Recently, clinician scientists from the International League Against Epilepsy (ILAE) Neuropsychology Task Force and the International Neuropsychological Society devised a taxonomy of cognitive disorders in epilepsy (known as *I*nternational *C*lassification for *Co*gnitive *D*isorders in *E*pilepsy, IC-CoDE) [11], which has been successfully applied in both temporal lobe epilepsy [12] and multiple sclerosis[13]. This taxonomy permits a harmonized approach to cognitive diagnostics across centers and batteries, because it focuses on cognitive constructs, not specific neuropsychological tests. The aims of this study were to: 1) adapt the IC-CoDE framework to examine cognitive dysfunction and diagnostic phenotypes in adults with post-COVID syndrome, 2) develop a complementary data-driven approach based on unsupervised machine-learning clustering algorithms to evaluate the optimal classification of patients according to cognitive function, 3) discover potential cognitive phenotypes in the post-COVID syndrome, and 4) establish the most meaningful cutoff points for the definition of cognitive impairment associated with COVID-19.

### 2. METHODS

### 2.1. Participants and study protocol

Four hundred and four patients with post-COVID syndrome at least three months after the acute infection were included in this study. All patients met the diagnostic criteria developed by WHO for the post-COVID condition [5] and reported new-onset subjective cognitive complaints after contracting COVID-19. The mean age was 48.6 ± 9.2 years and 80.2% were women. The main demographic characteristics are summarized in **Table 1**. Two centers participated in the present study: the Hospital Clinico San Carlos (HCSC) in Madrid, Spain; and the University of Oviedo (UO) in Oviedo, Spain. Both centers evaluated patients with cognitive dysfunction after COVID-19: HCSC clinically and UO only for research purposes. Neuropsychological tests used in each center are shown in **Supplementary Table 1**. Neuropsychological examinations were performed in person in HCSC and virtually in UO, in all cases by trained neuropsychologists.

**Table 1.** Demographic and clinical characteristics. Number (%) and mean ± Standard Deviation are shown.

| **Table 1.** Demographic and clinical characteristics. Number (%) and mean ± Standard Deviation are shown. | | | | |
| --- | --- | --- | --- | --- |
|  | HCSC<br>(n=157) | UO<br>(n=247) | X<sup>2</sup> or T (p-value) | Total<br>(n=404) |
| Age at time of<br>assessment<br>(years) | 50.18 ± 11.12 | 47.64 ± 7.61 | 2.51 (0.013) | 48.62 ±<br>9.20 |
| Years of<br>education | 14.56 ± 3.65 | 15.90 ± 3.20 | -3.88 (<0.001) | 15.30 ±<br>3.44 |
| Sex (% women) | 111 (70.7%) | 213 (86.23%) | 14.58 (<0.001) | 324 (80.2%) |
| Time since<br>COVID-19<br>onset (days) | 430.87 ± 195.73 | 518.75 ± 150.19 | -4.79 (<0.001) | 484.60 ±<br>174.46 |
| Arterial<br>hypertension | 39 (24.8%) | 23 (9.3%) | 17.81 (<0.001) | 62 (15.34%) |
| Diabetes | 21 (13.4%) | Not collected | - | - |
| Dyslipidemia | 45 (28.7%) | Not collected | - | - |
| Hospital<br>admission | 43 (27.4%) | 64 (25.9%) | 0.10 (0.742) | 107 (26.5%) |
| ICU admission | 11 (7.0%) | 6 (2.4%) | 4.98 (0.025) | 17 (4.2%) |

All patients had RT-PCR confirmation of COVID-19 infection, and other potential causes of cognitive dysfunction (e.g., any cognitive complaint before COVID-19, history of neurological or psychiatric disorders) beyond COVID-19 were considered exclusion criteria. Most of the neuropsychological tests in this study were co-normed in healthy adults from our setting[14-15]. Accordingly, raw scores for each test were converted to age-, education- and sex-adjusted scaled scores following these norms. For the other tests, normative data were also available from other studies[16-18].

### 2.2. Application of IC-CoDE definitions of impairment: IC-CoDi-COVID

Cognitive phenotypes were derived following the model proposed in the IC-CoDE seminal studies in epilepsy[11-12]. This model is based on five cognitive domains: language, memory, executive functioning, visuospatial abilities, and attention/processing speed. The neuropsychological tests selected by domain and center are shown in **Table 2**. These tests were selected following the recommendations from the IC-CoDE initiative, according to target cognitive domains, target abilities and desired test characteristics[11-12]. Tests are classified following two cutoffs: ≤1.0 SD and ≤1.5 standard deviations below the normative mean. The two most representative tests of each domain for each battery were selected. According to the model, a cognitive domain is impaired when both tests are below the established cutoff[11]. Then, the patients are categorized as 1) intact versus impaired cognition; 2) if impaired, single domain impairment; bi-domain impairment; or generalized (three or more cognitive domains impaired). Only patients with all the selected tests completed (n=391, 96.8%) were included in this analysis.

**Table 2.** Neuropsychological tests selected by domain and center

| **Table 2.** Neuropsychological tests selected by domain and center |  |  |
| --- | --- | --- |
|  | **HCSC** | **UO** |
| *Attention and processing speed* | SDMT<br>Stroop trial 1 | Brief Test of Attention<br>Stroop trial 1 |
| *Episodic memory* | FCSRT delayed total recall<br>ROCF recognition | TAVEC delayed recall with cues<br>ROCF delayed recall |
| *Executive function* | Digit span backwards<br>Stroop trial 3 | Digit span backwards<br>Stroop trial 3 |
| *Visuospatial* | JLO<br>VOSP (number location) | ROCF (copy)<br>WAIS-IV Matrix reasoning |
| *Language* | Boston Naming Test<br>Semantic fluency (animals) | Object naming<br>Semantic fluency (animals) |
| SDMT: Symbol Digit Modalities Test; FCSRT: Free and Cued Selective Reminding Test; JLO: Judgment Line Orientation;TAVEC: Test de Aprendizaje Verbal España-Complutense; ROCF: Rey-Osterrieth Complex Figure; VOSP: Visual Object and Space Perception Battery. |  |  |

### 2.3. Standard protocol approvals and patient consents

All procedures performed were in accordance with the ethical standards of the institutional research committee and with the 1964 Helsinki declaration and its later amendments. The local Research Ethics Committee approved the research protocol. All participants (or their legal representatives) gave written informed consent. No part of the study procedures or analyses was pre-registered in a time-stamped, institutional registry prior to the research being conducted, although decisions regarding design and analysis were decided a priori.

### 2.4. Statistical and machine learning analysis

Descriptive data are shown as frequency (percentage) and mean ± standard deviation. Chi-squared was used to compare qualitative variables. Two-independent samples t-test was used to evaluate differences in quantitative variables between the groups showing cognitive impairment or not. Findings were considered statistically significant when the p-value was <0.05 (two-tailed).

We applied Ward’s Hierarchical Agglomerative clustering method. This unsupervised machine learning algorithm analyzes the variance of clusters and is considered one of the most suitable clustering methods for quantitative variables. We used the adjusted scores of the cognitive tests selected from the two cohorts, and clustering analysis was performed in each dataset independently to demonstrate reliability and reproducibility of findings. The optimal number of clusters was defined according to the silhouettes scores, which measure how similar a data point is within a cluster (cohesion) compared to other clusters (separation). The silhouette coefficient value is between -1 and 1, where a score of 1 denotes the best fit, meaning that the data point is very compact within the cluster to which it belongs and far away from the other clusters, and values near 0 denote overlapping clusters. We also estimated the Dunn Index, a metric score that aims to quantify the compactness and variance of the clustering to evaluate the clustering algorithm. A higher Dunn Index indicates compact, well-separated clusters, while a lower index indicates less compact or not well-separated clusters. Clustered data requires a Dunn Index greater than 1. Other clustering algorithms (K-means, DBSCAN) were also investigated, but hierarchical clustering was preferred according to the specified parameters.

We also implemented Random Forest supervised classification models. The original dataset from each center was randomly divided into training (70%) and test (30%) sets to ensure the results’ reliability and reproducibility. The target was the classification between cognitively impaired (at least one cognitive domain) and cognitively intact according to the IC-CoDi-COVID criteria using the -1.0 SD and -1.5 SD cutoffs. A 5-fold cross-validation grid search was performed on the training set. Subsequently, the best model obtained was evaluated on the test set. Random forest models were used to rank the best cognitive tests from each center for the classification, considering the mean decrease in accuracy when a variable is removed from the model, and total increase in node purity.

## 3. RESULTS

### 3.1. Frequency of impairment on specific tests

The most frequent impairments occurred on the Stroop test and the verbal list learning tests (FCSRT and TAVEC). The most impaired score was the Stroop trial 1. All the percentages of patients with impaired test performance using both 1.0 SD and 1.5 SD cutoffs are shown in **Figure 1**.

**Figure 1.**
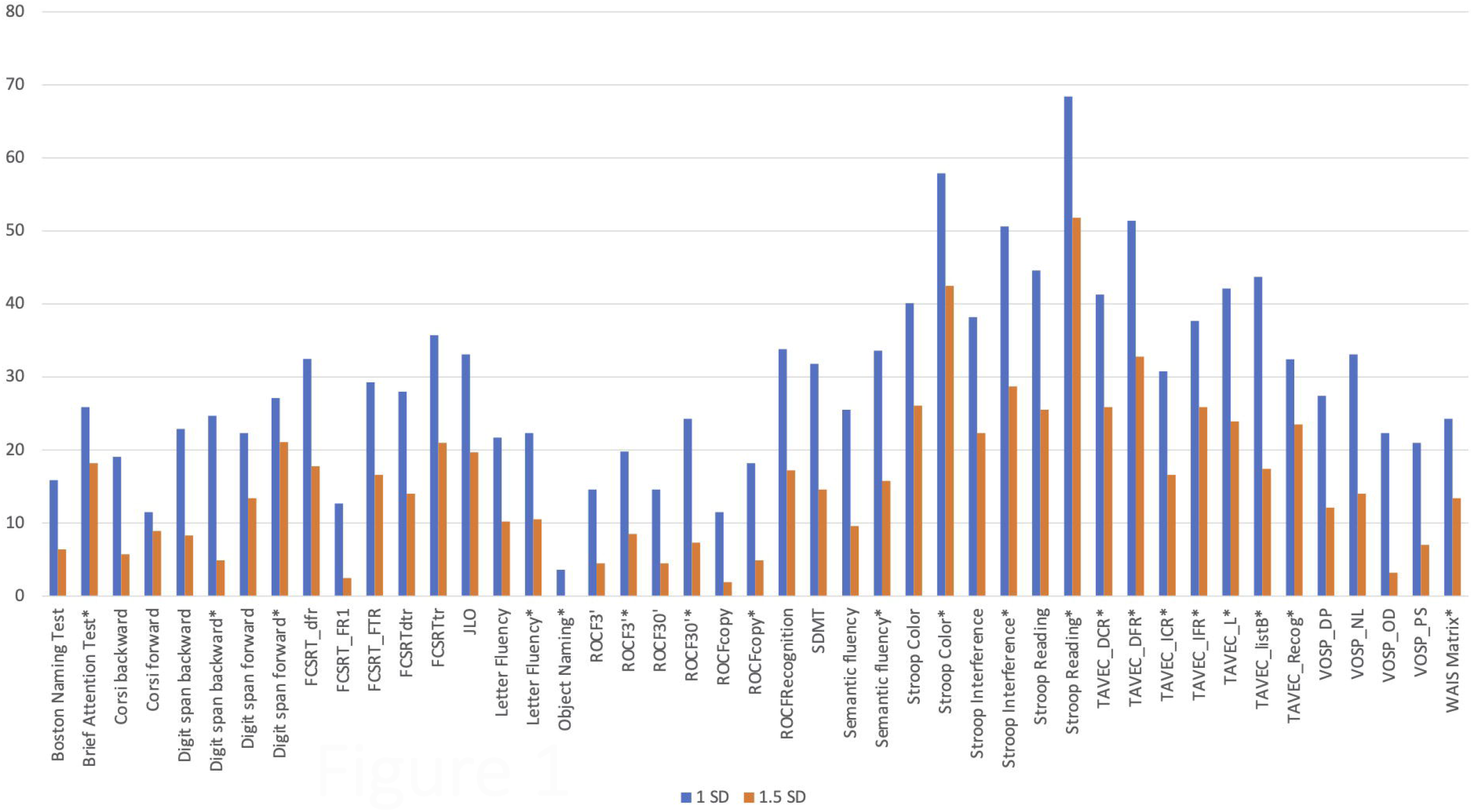
Percentage of impairment for each test according to -1.0 SD (blue) and -1.5 SD (orange) cutoffs. Tests are shown in alphabetical order. Scores with * were from the UO cohort. FCSRT: Free and Cued Selective Reminding Test (FR1: Free Recall trial 1; FTR: Free Total Recall; dtr: delayed total recall; tr: total recall); ROCF: Rey-Osterrieth Complex Figure (3’: recall at 3 minutes; 30’: recall at 30 minutes); SDMT: Symbol Digit Modalities Test; TAVEC: Test de Aprendizaje Verbal España-Complutense (DCR: Delayed Cued Recall; DFT: Delayed Free Recall; ICR: immediate cued recall; IFR: immediate free recall; L: Learning; Recog: Recognition); VOSP: Visual Object and Space Perception Battery (DP: discrimination of position; NL: number location; OD: object decision; PS: progressive silhouettes).

### 3.2. Cognitive phenotypes using IC-CoDi-COVID criteria

Using the 1.0 SD cutoff, 230 patients (58.8% of the sample) were classified as cognitively intact and 161 (41.2%) as cognitively impaired in at least one cognitive domain. Single-domain impairment was observed in 86 (22.0%), two domains were impaired in 40 (10.2%), with generalized impairment in 35 (9.0%) cases. The most frequent cognitive domain impaired was attention/processing speed (93 patients, 23.3%), followed by episodic memory (72, 18.0%), executive function (64, 16.1%), visuospatial function (47, 11.8%), and language (20, 5.0%) (**Figure 2A)**. In patients with single-domain impairment, the most common domain was attention/processing speed, followed by episodic memory and executive function (**Figure 3A)**. In patients with bi-domain impairment, the most frequent combinations were attention/processing speed and executive dysfunction, followed by attention/processing speed associated with deficits in episodic memory or visuospatial skills. In patients showing multi-domain impairment, the most frequent combinations included attention/processing speed, episodic memory, and executive functioning (**Figure 4A**).

**Figure 2.**
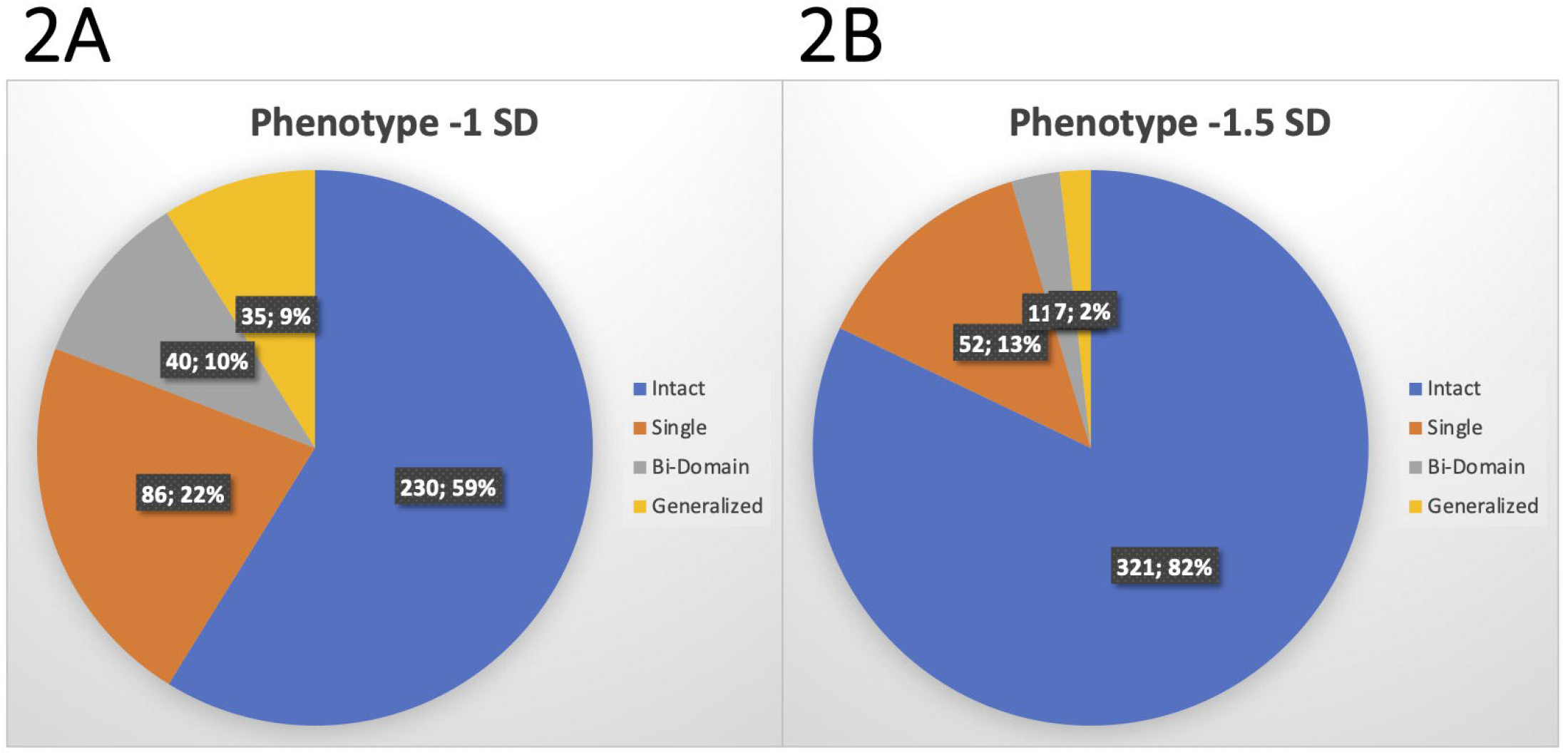
Cognitive phenotyping according to the IC-CoDi-COVID classification. **A**: Using -1.0 SD cutoff; **B**: Using -1.5 SD cutoff.

**Figure 3.**
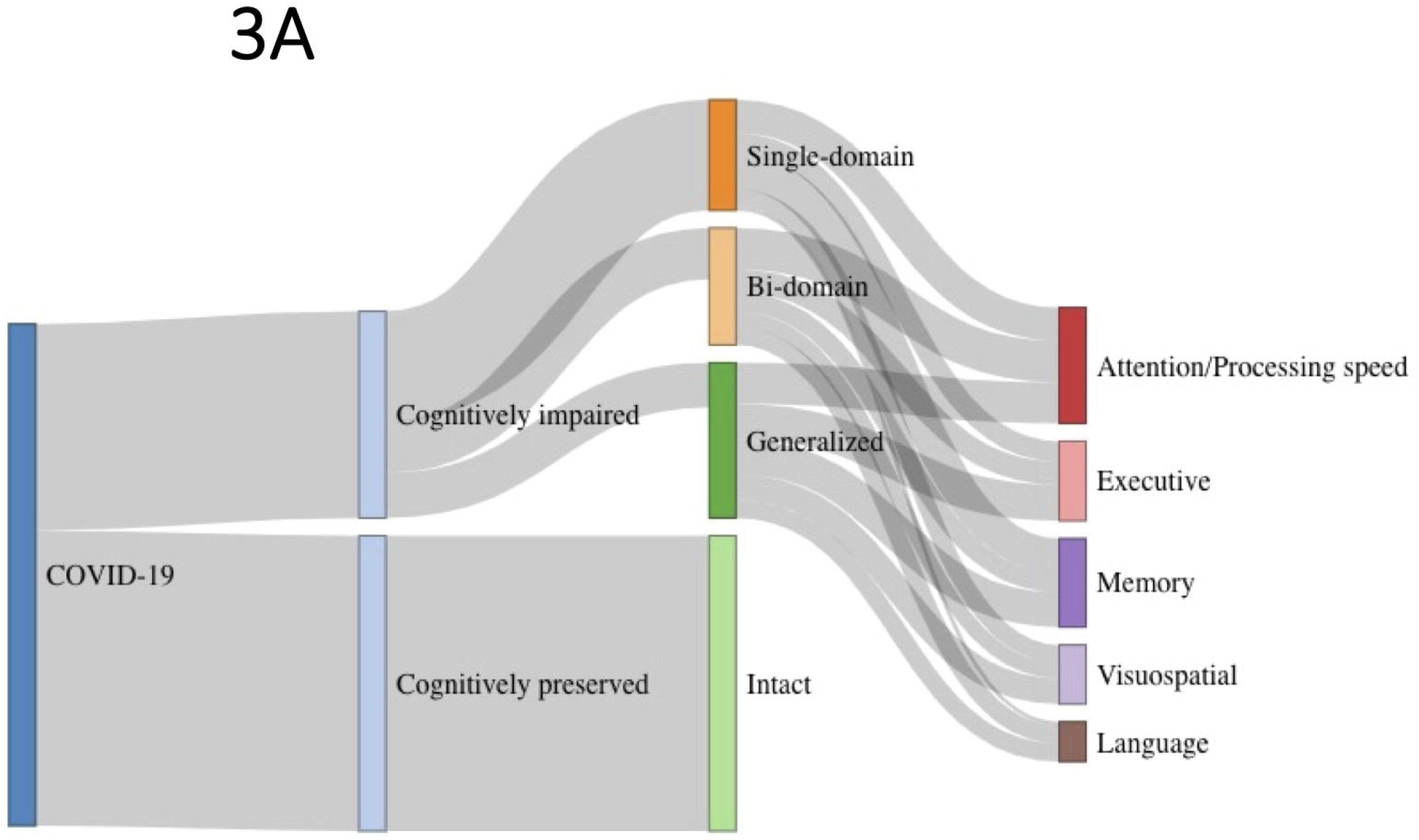
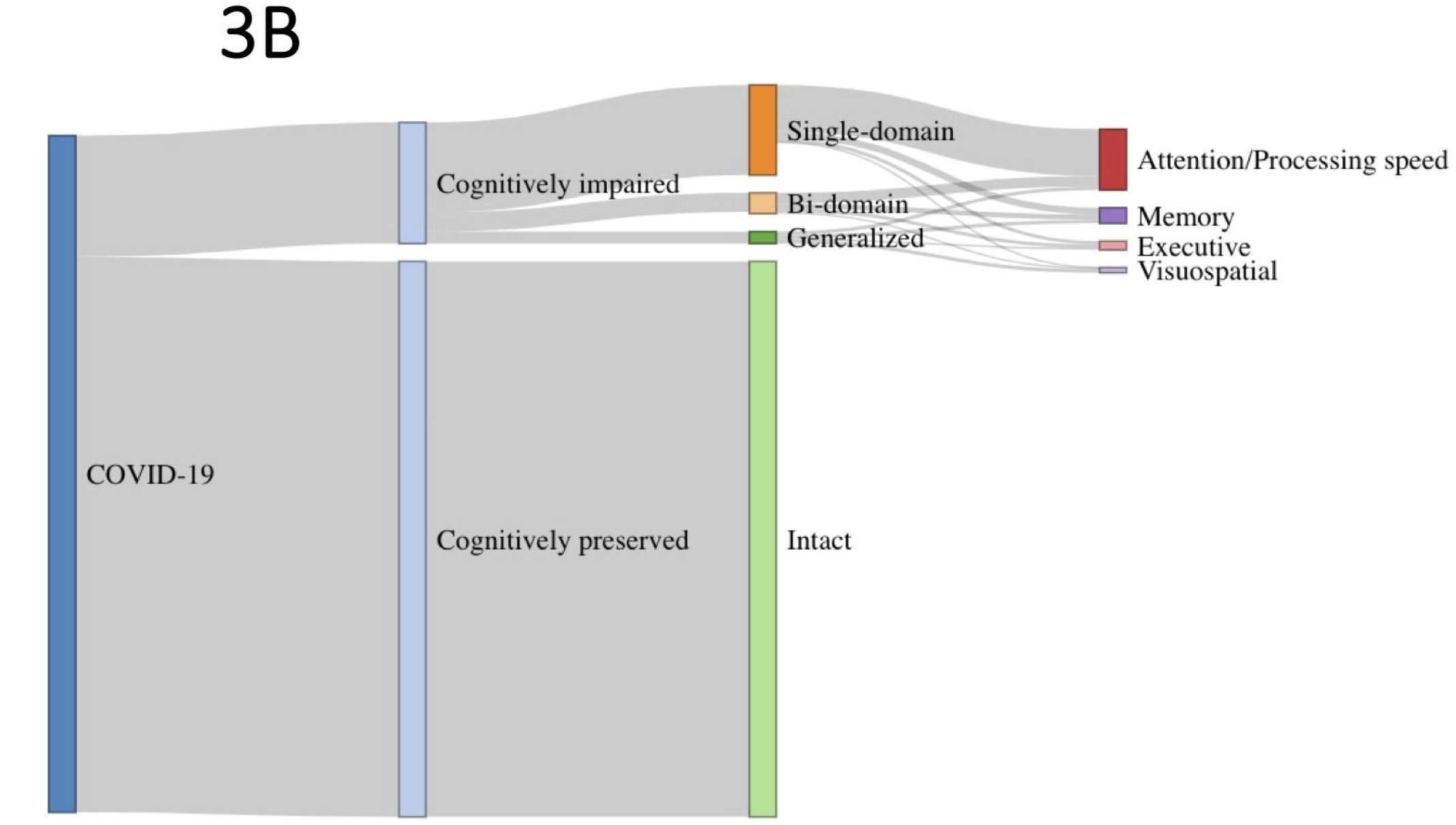
Sankey diagrams showing the flow of patients according to the IC-CoDi-COVID classification and 5-cognitive domains. **A**: Using -1.0SD cutoff; **B:** Using -1.5 SD cutoff.

**Figure 4.**
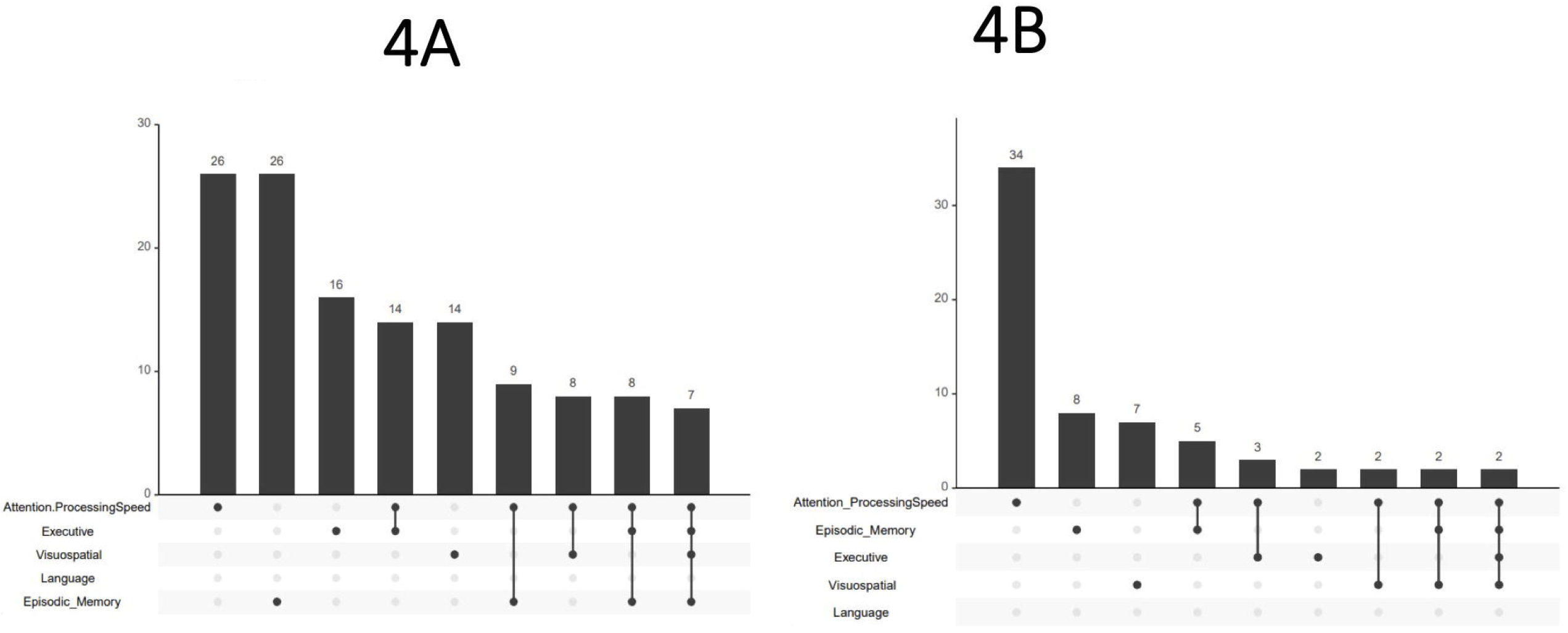
Plot of intersections between cognitive domains impaired. **A**: Using -1.0 SD cutoff; **B:** Using -1.5 SD cutoff. The upper bar chart shows the number of cases displaying the intersections. Connected dots on the bottom panel mean which cognitive domains are considered for each intersection.

Using the 1.5 SD cutoff, 321 patients (79.5% of the sample) were classified as cognitively intact, and 70 (17.3%) as cognitively impaired in at least one cognitive domain. Single-domain impairment was observed in 52 patients (19.9%), two domains were impaired in 11 (2.7%), and the phenotype revealed a generalized impairment in 7 (1.7%) cases (**Figure 2B)**. The most frequent cognitive domain impaired was attention/processing speed, followed by episodic memory, executive function, visuospatial function, and language. In patients with single-domain impairment, the most common domain altered was attention/processing speed, followed by memory and executive function (**Figure 3B B**). The most frequent combinations included attention/processing speed along other cognitive domains (**Figure 4B**).

Cognitive phenotypes, separately by Center, are shown in **Supplementary Figures 1 and 2**. There were no statistically significant differences in the cognitive phenotypes (intact, single-domain, bi-domain and generalized impairment) between the two centers (X^2^=4.028,p=0.259 for -1.0 SD cutoff; X^2^=3.226,p=0.358 for -1.5 SD).

### 3.3. Association between cognitive phenotypes and demographic and hospitalization

Patients displaying cognitive impairment at -1.0 SD were younger (46.65±9.08 vs 50.05±9.05,T=3.64,p<0.001) and showed lower levels of education (14.99±3.58 vs 15.73±3.30, T=2.07,p=0.039). There was no association between cognitive phenotype and interval from acute onset to neuropsychological assessment (483.09±170.95 days for the cognitively intact group vs 489.82±180.25 for the cognitive impaired group, T=- 0.375,p=0.708). Regarding sex, 58.4% of men and 58.9% of women were classified as cognitively intact (X^2^=0.006,p=0.939). Cognitive phenotypes did not differ between patients who had been hospitalized or admitted to the ICU and those who had not (29.2% of those showing cognitive impairment vs 23.7% of cognitively intact were hospitalized,X^2^=1.614,p=0.204; 2.5% of cognitively impaired vs 4.3% of cognitively intact required ICU,X^2^=0.952,p=0.329)

Patients showing cognitive impairment at -1.5 SD were younger (45.76±9.44 vs. 49.28±9.04,T=2.929,p=0.004) and showed a trend to have lower levels of formal education (14.77±3.66 vs. 15.57±3.37, T=0.102,p=0.085). There were no differences in the time since COVID-19 onset between both groups (484.10±172.37 in cognitively intact vs 493.94±185.82 in cognitively impaired, T=-0.427,p=0.670). Sex distribution was also similar between both groups (81.8% and 82.2% of men and women were regarded as cognitively intact, respectively, X^2^=0.005,p=0.943). The frequency of hospitalization was 77 (24.0%) in the cognitively intact group and 24 (34.3%) in the group displaying cognitive impairment (X^2^=3.181, p=0.074). Ten patients (3.1% of cases) with no cognitive impairment and four patients (5.7%) with cognitive impairment were admitted to the ICU (X^2^=1.124,p=0.289).

### 3.4. Machine learning classification Hierarchical clustering

For the HCSC cohort, the optimal number of clusters was 2 (silhouette score 0.190, Dunn index 0.188). The first cluster included 73 cases and comprised mainly of patients showing cognitive impairment (24.73% of the intact group, 56.66% of single-domain, 93.33% of bi-domain and 100% of generalized impairment according to -1.0SD cutoff). In the comparison with the classification using -1.5 SD cutoff, this cluster comprised 35.93% of patients with intact cognition, 89.47% of patients showing single-domain impairment, and 100% of bi-domain and generalized impairment. The second cluster involved 84 patients and were mainly cases with intact cognition (75.26% of the intact group, 43% of the single-domain, 6.66% of bi-domain, and 0% of generalized impairment groups following the -1SD cutoff; and 64.06% of the intact group, 10.52% of single-domain, and 0% of bi-domain and generalized impairment groups according to the -1.5SD cutoff)

For the UO cohort, the optimal number of clusters was also 2 (silhouette score 0.180, Dunn index 0.168). A first cluster involved 140 patients and included patients with cognitive impairment (87% of single-domain, 100% of bi-domain and 100% of generalized domain according to -1.0SD cutoff). 36.4% of the intact group were classified in the first cluster. The second cluster included 94 patients, and were mainly patients within the intact group (63.5% of this group fell in this cluster, 12.5% of the single-domain). When comparing clustering analysis with -1.5SD classification, the first cluster included 52.3% of patients classified as intact, 93.93% of single-domain, and 100% of bi-domain and generalized impairment. Patients in the second cluster comprised mainly intact cases (97.87% of patients in the second cluster were classified as intact; however, 47.66% of patients in the intact group belonged to the second cluster).

### 3.5. Random Forest classification

The models obtained a high level of accuracy for both cohorts and the -1.0 SD and -1.5 SD cutoffs. The models’ evaluation metrics (accuracy, precision, recall, F1-score and area under the curve) are shown in **Supplementary Table 2**. The most important features were generally tests associated with attention/processing speed, episodic memory, and executive function. ROC curves and feature importance are shown in **Figure 5**.

**Figure 5.**
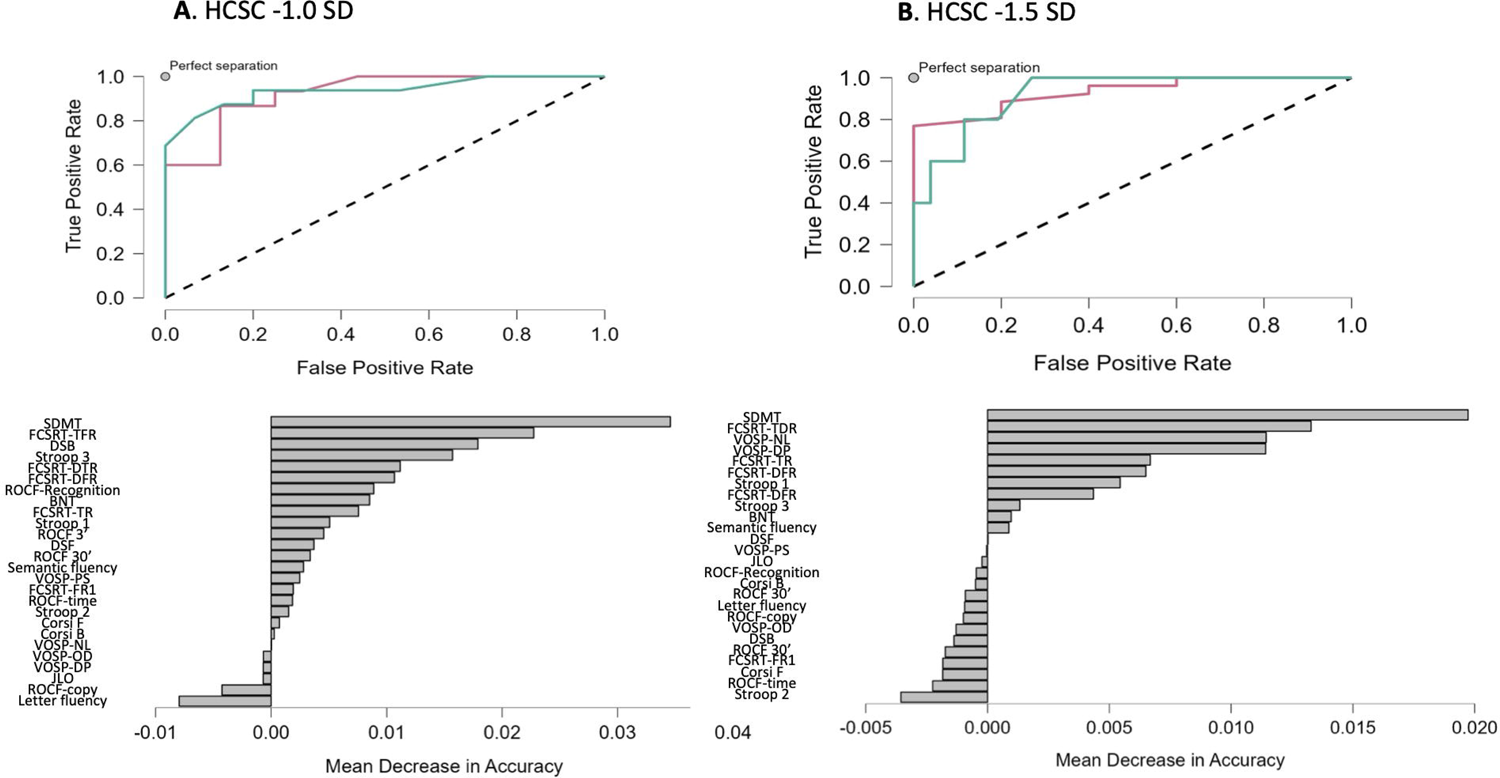
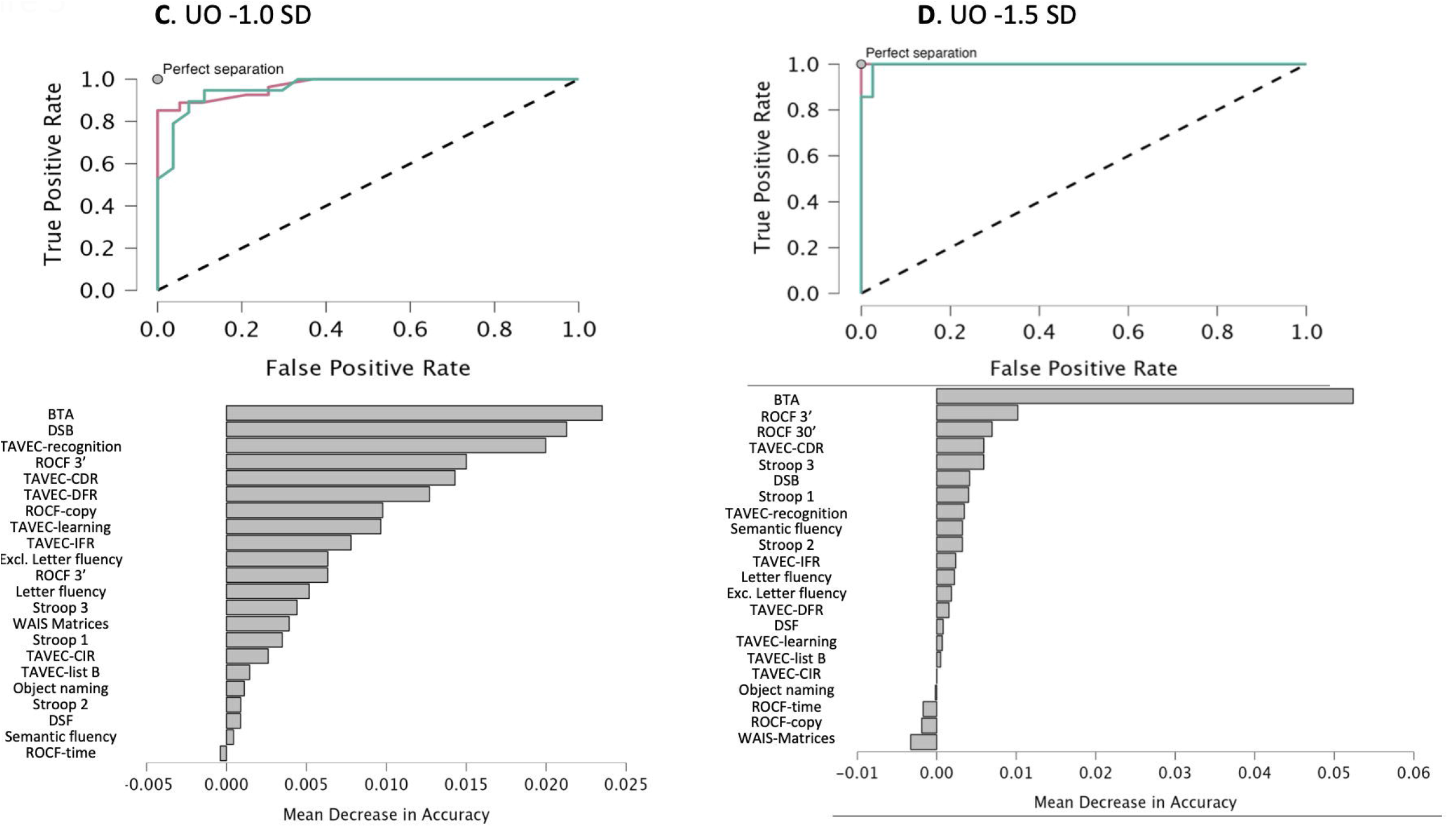
Random forest for classification between patients cognitively impaired and cognitively intact according to IC-CoDi-COVID approach. ROC curves and mean decrease in accuracy are shown for the two centers. **A**: HCSC, with -1.0 SD as cutoff; **B**: HCSC using -1.5 SD as cutoff; **C:** UO with -1.0 SD as cutoff; and **D**: UO with -1.5 SD as cutoff. Green line: cognitively impaired group; Red line: cognitively intact group. BNT: Boston Naming Test; BTA: Brief Test of Attention; DSB: digit span backward; DSF: digit span forward; FCSRT: Free and Cued Selective Reminding Test (FR1: Free Recall trial 1; FTR: Free Total Recall; dtr: delayed total recall; tr: total recall); JLO: Judgment Line Orientation; ROCF: Rey-Osterrieth Complex Figure (3’: recall at 3 minutes; 30’: recall at 30 minutes); SDMT: Symbol Digit Modalities Test; TAVEC: Test de Aprendizaje Verbal España-Complutense (DCR: Delayed Cued Recall; DFT: Delayed Free Recall; ICR: immediate cued recall; IFR: immediate free recall; L: Learning; Recog: Recognition); VOSP: Visual Object and Space Perception Battery (DP: discrimination of position; NL: number location; OD: object decision; PS: progressive silhouettes).

## 4. DISCUSSION

In this study, we adapted a classification system for cognitive phenotyping from other neurological disorders to the post-COVID syndrome. This approach was applied to two large cohorts of patients with the post-COVID syndrome that underwent a comprehensive neuropsychological assessment for clinical purposes. Interestingly, the prevalence of cognitive deficits was very similar in both cohorts. Prevalence of cognitive impairment was 41.2% considering the -1 SD cutoff, and 17.3% with -1.5 SD. These findings confirm in a large sample the presence of cognitive deficits in patients with the post-COVID syndrome, which mainly involve attention/processing speed, executive functioning, and episodic memory. In addition, this cognitive phenotype is consistent across centers and with two different impairment thresholds. Overall, these findings suggest that the taxonomy was robust despite some variation in the specific tests used and in the characteristics of the population. This is consistent with prior reports in epilepsy and multiple sclerosis [12-13] and the main goal of IC-CODE initiative.

One of the most striking findings in our study is the presence of two clusters of patients according to neuropsychological assessment results. The optimal 2-cluster solution was replicated in both cohorts, confirming the validity of the results. However, the different metrics (Dunn Index, silhouettes) show a weak level of separation between clusters. This suggests that cognitive phenotypes are relatively homogeneous and that post-COVID syndrome patients exhibit a common cognitive profile. In this regard, the separation into two clusters was due to the severity of cognitive dysfunction rather than the overall cognitive profiles. Therefore, the unsupervised method could not find distant and compact clusters associated with cognitive profiles. However, the application of the taxonomy of IC-CoDi-COVID may identify specific cognitive groups, which could be clinically useful and supports the application of the proposed taxonomy. Future research is needed to link this categorization with biomarkers including neuroimaging metrics.

On the one hand, this supports the consistency of the cognitive impairment post-COVID syndrome, which has a relatively characteristic cognitive profile characterized by attention/processing speed deficits with or without associated episodic memory and executive function deficits. This has important implications for the differential diagnosis because the identification of patients after COVID-19 displaying other cognitive profiles with a predominant impairment of other cognitive domains beyond attention/processing speed (e.g., visuospatial, language) should suggest alternative diagnoses[19]. On the other hand, clustering analysis may be helpful in determining the optimal cutoff points for patient classification following an unsupervised strategy. In this regard, the comparison between IC-CoDi-COVID classification and clustering analysis suggested that using a stricter criterion of -1.5 SD, patients with one or more cognitive domains impaired should be classified as cognitively impaired. However, in this case, a relatively substantial percentage of patients that should be considered impaired according to the clustering analysis fall into the “intact” group according to the IC-CoDi-COVID system. Thus, this cutoff should be regarded as having high specificity but low sensitivity. Conversely, when using the -1.0 SD cutoff criterion, two or more cognitive domains impaired are almost always included in the cognitively impaired cluster. When one cognitive domain is impaired, the probability of being included in the impaired cluster is higher, but there is some degree of uncertainty. In contrast, up to 24-36% of patients within the “intact” phenotype are included in the impaired cluster, which represents a lower percentage than using the stricter criterion. Overall, results from clustering analysis suggest that the impairment of one cognitive domain with -1.5 SD cutoff or two cognitive domains with -1.0 SD cutoff should be considered as cognitively impaired in the setting of the post-COVID syndrome. The observation of one cognitive domain impaired using -1.0 SD should probably also be considered as cognitively impaired, although it would require more careful and detailed analysis. The coherence between clustering analysis and IC-CoDi-COVID classification further validates this latter approach from an unsupervised perspective[20].

The consistency of the cognitive phenotype could suggest a common pathophysiology for cognitive dysfunction in patients with the post-COVID syndrome due to the similarities in the cognitive profile. In this regard, these criteria for cognitive dysfunction could also be useful for future studies focused on the identification of novel biomarkers and neuroimaging analysis[21-23].

Furthermore, we implemented machine-learning algorithms to detect cognitive dysfunction in post-COVID syndrome. In recent years, this approach has gained interest in improving the interpretation of neuropsychological assessment and test selection to reduce the length of the examinations and generalize cognitive assessments in clinical practice[24]. In this regard, we obtained high accuracy and F1-scores, and the most meaningful tests in the classification confirmed the importance of assessing attention/processing speed and episodic memory in these patients.

Another interesting finding of our study is the association between cognitive impairment and some demographic factors. Specifically, patients displaying cognitive impairment according to the criteria were younger and less educated. On the one hand, this suggests a special vulnerability to post-COVID syndrome in the young population, which could have implications from a pathophysiological perspective. On the other hand, these findings confirm the role of the cognitive reserve, as has been observed in several other disorders[25]. Regarding sex, we did not find significant differences in the presence of cognitive impairment, although the frequency of females in both cohorts was elevated. Conversely, our study did not find significant associations with hospitalization or ICU admission. Overall, this suggests that cognitive impairment could be independent of the severity in the acute phase, and thus, cognitive symptoms are not merely sequelae of complications during the acute phase of the disease.

The mean time between the acute onset of COVID-19 and the neuropsychological assessment was over 16 months. This is a longer time compared with other studies [26]. Although we cannot draw conclusions about the outcome of cognitive dysfunction in post-COVID syndrome, the detection of cognitive impairment in patients and the absence of differences in the time between patients cognitively intact and cognitively impaired suggest that this condition could persist for at least 1-2 years. Future studies should evaluate the outcome of cognitive dysfunction using a longitudinal design. In addition, potential differences between SARS-CoV-2 variants in the induction of cognitive consequences and the preventive effect of vaccination deserve further investigation.

Our study has some limitations that should be considered. First, both cohorts are from the same country. Replication studies in other settings should be performed to confirm the generalizability of this approach. Due to the positive experience in epilepsy and multiple sclerosis fields[12-13], the application to other cultures and languages should be feasible. Second, one of the batteries was administered virtually, but normative data were obtained from previous studies performing in-person neuropsychological assessments. However, considering the consistency with the findings from the cohort, in which all patients were evaluated in person, these results could indirectly suggest the viability of virtual neuropsychological assessment in this population. Third, we did not include a control group of patients who contracted COVID-19 but did not report cognitive complaints.

In conclusion, we propose a harmonization of the criteria for the definition of cognitive impairment in the post-COVID syndrome, based on the framework developed by IC-CoDE and supported by unsupervised machine learning algorithms. Cognitive issues are among the most frequent symptoms in the post-COVID syndrome, and, in contrast to other symptoms, they may be accurately quantified with neuropsychological assessment. This study provides a taxonomy to classify patients according to their cognitive status objectively. This approach may be generalized to other cognitive batteries and samples of patients with post-COVID syndrome and is a first step to developing international and unified criteria for the definition of cognitive impairment in post-COVID syndrome.

## Data Availability

All data produced in the present study are available upon reasonable request to the authors

## Acknowledgements

We are grateful to all the participants for their interest in the study. We thank the “LongCovid ACTS association” and “Colectivo COVID-19 persistente Madrid” for their collaboration. We also appreciate the help of the other IC-CoDE members (Carrie McDonald, Sallie Baxendale, William Barr, David Loring, Cady Block, Erik Hessen) and IC-CoDi-MS (Laura M Hancock, Rachel Galioto). We also thank Prof. Fernando Cuetos for his support and advice with this project.

## Ethics approval

This study was approved by the Ethics and Research Committee from HCSC (code 20/633-E) and UO (code CEImPA 2021.487) and was performed according to the Declaration of Helsinki and its later amendments.

## Availability of data and material

The datasets generated and analyzed are available from the corresponding author on request.

## Supplementary Material

**Supplementary Figure 1**. Cognitive phenotyping according to the IC-CoDi-COVID classification using -1.0 SD cutoff for each center. Left: HCSC; Right: UO.

**Supplementary Figure 2**. Cognitive phenotyping according to the IC-CoDi-COVID classification using -1.5 SD cutoff for each center. Left: HCSC; Right: UO.

**Supplementary Table 1.** List of all the tests administered in each center.

**Supplementary Table 2.** Evaluation metrics of Random Forest Models for the classification between patients cognitively impaired (at least one domain below the cutoff) and cognitively preserved (intact cognition), according to the IC-CoDI-COVID system

**Supplementary Table 1.** Neuropsychological tests included in the batteries of each center.

|  |  |
| --- | --- |
| HCSC | UO |
| Digit span forward and backward | Digit span forward and backwards |
| Corsi block-tapping test forward and backward | - |
| Symbol Digit Modalities Test | Brief Test of Attention |
| Stroop Color-Word Interference test (word reading, color naming, and interference), | Stroop Color-Word Interference test (word reading, color naming, and interference) |
| Free and Cued Selective Reminding Test | TAVEC (a Spanish adaptation of the California Verbal Learning Test) |
| Rey-Osterrieth Complex Figure (ROCF) (copy and recall at 3 and 30 minutes and recognition), | ROCF (copy and recall at 3 and 30 minutes) |
| Visual Object and Space Perception Battery (discrimination of position, number location, object decision, and progressive silhouettes). | WAIS-IV Matrix reasoning |
| Verbal fluency (animals and words beginning with "p") | Verbal fluency (animals, words beginning with "p", and excluded letter) |
| Boston Naming Test | Object naming task |
| Judgement of Line Orientation |  |

**Supplementary Table 2.**
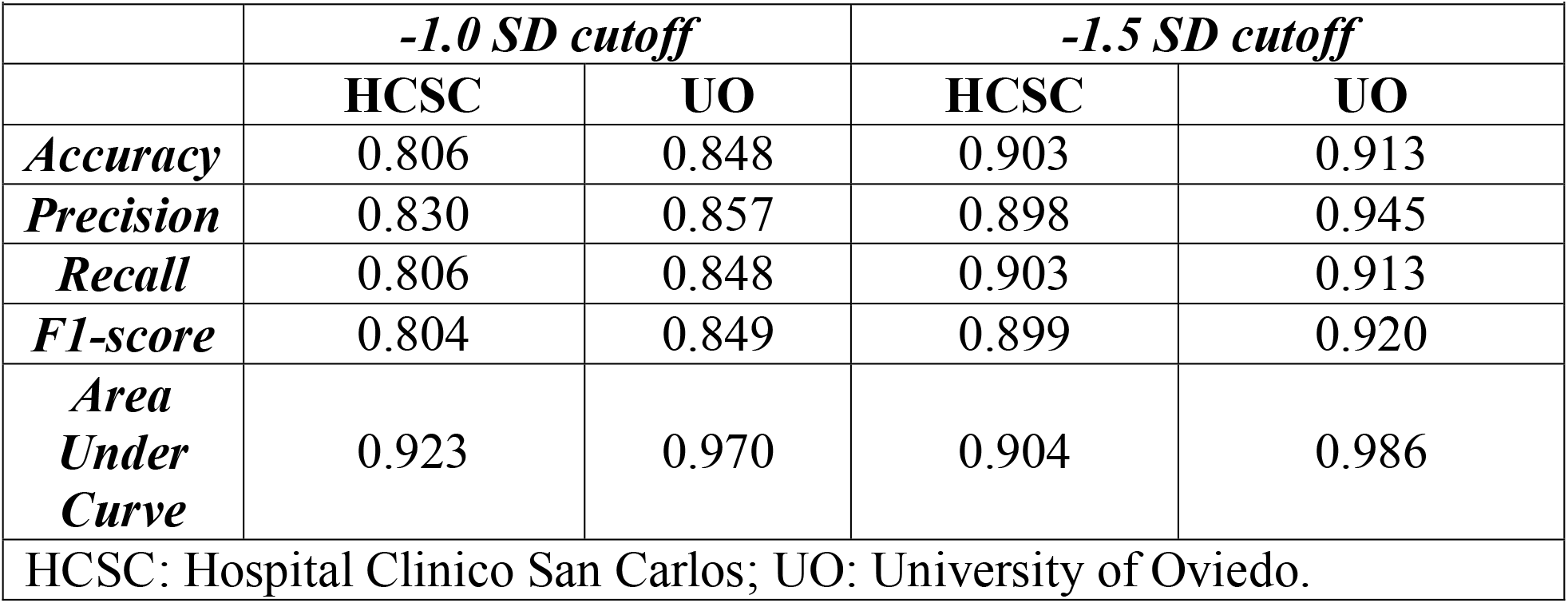
Evaluation metrics of Random Forest Models for the classification between patients cognitively impaired (at least one domain below the cutoff) and cognitively preserved (intact cognition), according to the IC-CoDI-COVID system

|  |  |  |  |  |
| --- | --- | --- | --- | --- |
|  | ***-1.0 SD cutoff*** |  | ***-1.5 SD cutoff*** |  |
|  | **HCSC** | **UO** | **HCSC** | **UO** |
| ***Accuracy*** | 0.806 | 0.848 | 0.903 | 0.913 |
| ***Precision*** | 0.830 | 0.857 | 0.898 | 0.945 |
| ***Recall*** | 0.806 | 0.848 | 0.903 | 0.913 |
| ***F1-score*** | 0.804 | 0.849 | 0.899 | 0.920 |
| ***Area Under Curve*** | 0.923 | 0.970 | 0.904 | 0.986 |
| HCSC: Hospital Clinico San Carlos; UO: University of Oviedo. |  |  |  |  |

